# Development and external validation of a diagnostic model for periprocedural bradycardia during primary percutaneous coronary intervention

**DOI:** 10.1101/2020.06.26.20140558

**Authors:** Yong Li, Shuzheng Lyu

**Affiliations:** Emergency and Critical Care Center, Beijing Anzhen Hospital, Capital Medical University, Beijing 100029, China; Department of Cardiology, Beijing Anzhen Hospital,Capital Medical University, Beijing100029, China

**Keywords:** coronary disease, ST elevation myocardial infarction, Bradycardia, Percutaneous coronary intervention, Nomogram

## Abstract

**Background:** Periprocedural bradycardia weaks the benefit of primary percutaneous coronary intervention (PPCI) and has deleterious effects on organ perfusion of patients with acute ST elevation myocardial infarction (STEMI).

**Objective:** To develop and externally validate a diagnostic model of periprocedural bradycardia..

**Methods:** Design: Multivariable logistic regression of a cohort of acute STEMI patients. Setting: Emergency department ward of a university hospital. Participants: Diagnostic model development: Totally 1820 acute STEMI patients who were consecutively treated with PPCI from November 2007 to December 2015 in Beijing Anzhen Hospital, Capital Medical University. External validation: Totally 716 acute STEMI patients who were treated with PPCI from January 2016 to June 2018 in Beijing Anzhen Hospital, Capital Medical University. Outcomes:Periprocedural bradycardia during PPCI. Periprocedural bradycardia was defined as preoperative heart rate ≥ 50 times / min, intraoperative heart rate <50 times / min persistent or transient.

**Results:** Totally 332 (18.2%)patients presented periprocedural bradycardia in the development dataset and 102 (14.2%) patients presented periprocedural bradycardia in the validation dataset. The strongest predictors of periprocedural bradycardia were intra-procedural hypotension, the culprit vessel was not left anterior descending (LAD), using thrombus aspiration devices during procedure, sex, history of coronary artery disease, total occlusion of culprit vessel, and no-reflow. We developed a diagnostic model of periprocedural bradycardia.The area under the receiver operating characteristic(ROC) curve(AUC) was 0.8384± 0.0122 in the development set. We constructed a nomogram based on predictors of periprocedural bradycardia using the development database. The AUC was 0.8437±0.0203 in the validation set. Discrimination, calibration, and decision curve analysis were satisfactory.

**Conclusions:** We developed and externally validated a diagnostic model of periprocedural bradycardia during PPCI.

We registered this study with WHO International Clinical Trials Registry Platform(ICTRP). Registration number: ChiCTR1900023214.

Registered Date :16 May 2019. http://www.chictr.org.cn/edit.aspx?pid=39087&htm=4

## Background

Periprocedural bradycardia weaks the benefit of primary percutaneous coronary intervention (PPCI) and has deleterious effects on organ perfusion of patients with acute ST elevation myocardial infarction (STEMI). ^[1]^ We want to develop and externally validate a diagnostic model of periprocedural bradycardia.

## Methods

We followed the methods of Li et al. 2019 ^[2]^and Li. 2020 ^[3]^.

We followed the methods of Transparent Reporting of a multivariable prediction model for Individual Prognosis Or Diagnosis (TRIPOD). ^[4]^We used type 2b of prediction model studies covered by the TRIPOD statement. We split the data nonrandomly by time into 2 groups: one to develop the prediction model and one to evaluate its predictive performance. ^[4]^Type 2b was referred to as “external validation studies”. ^[4]^

The derivation cohort was 1820 patients with acute STEMI presenting within 12 hours from the symptom onset who were consecutively treated with PPCI between November 2007 and December 2015 in Beijing Anzhen Hospital, Capital Medical University.

The validation cohort was 716 patients we recruited on the same basis between January 2016 and June 2018 in Beijing Anzhen Hospital, Capital Medical University.

Inclusion criteria: STEMI patients presenting within 12 hours from the symptom onset who were treated with PPCI. We established the diagnosis of AMI and STEMI base on fourth universal definition of myocardial infarction. ^[5]^

Exclusion criteria: 1. patients received thrombolysis ; 2. patients received bivalirudin.

Prior to emergency angiography, all patients received 300mg of aspirin,300 to 600 mg of clopidogrel or 180 mg of ticagrelor, and unfractionated heparin.

It was a retrospective analysis and informed consent was waived by Ethics Committee of Beijing Anzhen Hospital Capital Medical University.

Outcome of interest was periprocedural bradycardia. Periprocedural bradycardia was defined as preoperative heart rate ≥ 50 times/min, intraoperative heart rate <50 times / min persistent or transient. ^[1]^ Preoperative heart rate was based on the medical record, intraoperative heart rate was based on the operation record. The presence or absence of periprocedural bradycardia was decided blinded to the predictor variables and based on consensus and recorded on the operation record.

We selected 10 predictor variables for inclusion in our prediction rule according to clinical relevance and the results of baseline descriptive statistics. Ten potential candidate variables were no-reflow, age, sex, history of hypertension, history of diabetes, history of coronary artery disease, culprit vessel site, total occlusion of culprit vessel, intra-procedural hypotension (pre-procedural systolic blood pressure was> 90mmHg, intra-procedural systolic blood pressure less than or equal to 90 mmHg persistent or transient^[6]^), and using thrombus aspiration devices during procedure.

We used angiographic criteria for the diagnosis of no-reflow. ^[7]^We defined no-reflow as Thrombolysis In Myocardial Infarction flow grade (TIMI)< 3 after successful mechanical opening of the infarct-related arterie with a guide wire, a balloon, or a stent, not the end of the PPCI process. ^[8–10]^

Pre-procedural blood pressure, age, sex, history of hypertension, history of diabetes, and history of coronary artery disease were based on the medical record. No-reflow, culprit vessel site, total occlusion of culprit vessel, using thrombus aspiration devices during procedure, and intra-procedural hypotension were based on the operation record.

Some had suggested having at least 10 events per candidate variable for the derivation of a model and at least 100 events for validation studies. ^[4]^Our sample and the number of events exceeds all approaches for determining samples sizes and therefore was expected to provide estimates that were very robust. ^[4]^

To ensure reliability of data, we excluded patients who had missing information on key predictors: no-reflow, the culprit vessel was left anterior descending(LAD), using thrombus aspiration devices during procedure, sex, history of coronary artery disease, total occlusion of culprit vessel, and intra-procedural hypotension.

### Statistical analysi

We followed the methods of Li et al. 2019 ^[2]^and Li. 2020 ^[3]^.

We used univariable and multivariable logistic regression models to identify the correlates of periprocedural bradycardia during PPCI. We used the Akaike information criterion (AIC)and Bayesian information criterion(BIC)to select predictors; it accounts for model fit while penalizing for the number of parameters being estimated and corresponds to using α =0.157. ^[4]^

We assessed the predictive performance of the diagnostic model in the validation data sets by examining measures of discrimination, calibration, and decision curve analysis (DCA).

Statistical analyses were performed with STATA version 15.1 (StataCorp, College Station, TX) and R version 4.0.0(R Development Core Team; http://www.r-project.org) and the RMS package developed by Harrell (Harrell et al). All tests were two-sided and a P value <0.05 was considered statistically significant.

## Results

Totally 332 patients had periprocedural bradycardia (periprocedural bradycardia group) and 1488 patients had no periprocedural bradycardia (control group) in the development data set. Baseline characteristics of the patients were shown in Table 1.

**Table 1.** Demographic, clinical, and angiographic characteristics of patients with periprocedural bradycardia and control group during PPCI

| Characteristic<br>[lower limit, upper limit] | Development data sets |  |  |  | Validation data sets |  |  |  |
| --- | --- | --- | --- | --- | --- | --- | --- | --- |
|  | Total<br>(1820) | Periprocedural<br>bradycardia<br>group(n =332) | Control<br>group<br>(n=1488) | P<br>value | Total<br>(716) | Periprocedural<br>bradycardia<br>group(n =102) | Control<br>group<br>(n=614) | P<br>value |
| Age,years[23,80] | 57±11 | 58±11 | 57±11 | 0.029 | 57±11 | 59±11 | 56±11 | 0.043 |
| Man n(%) 0=No, 1=Yes | 1507(82.8) | 255(76.8) | 1252(84.1) | 0.001 | 606(84.6) | 83(81.4) | 523(85.2) | 0.325 |
| History of coronary artery disease<br>n(%)0=No, 1=Yes | 967(53.1) | 189(57) | 778(52.3) | 0.126 | 282(39.4) | 31(30.4) | 251(40.9) | 0.046 |
| History of hypertension n(%)0=No, 1=Yes | 939(51.6) | 172(51.8) | 767(51.5) | 0.931 | 387(54.1) | 53(52) | 334(54.4) | 0.648 |
| History of diabetes n(%)0=No, 1=Yes | 449(24.7) | 76(22.9) | 373(25.1) | 0.406 | 203(28.4) | 25(24.5) | 178(29) | 0.353 |
| Culprit vessel site n(%) 0=No, 1=Yes |  |  |  |  |  |  |  |  |
| Left anterior descending<br>coronary artery | 880(48.4) | 54(16.3) | 826(55.5) | <0.001 | 330(46.1) | 8(7.8) | 322(52.4) | <0.001 |
| Left circumflex coronary artery | 215(11.8) | 47(14.2) | 168(11.3) | 0.144 | 88(12.3) | 12(11.8) | 76(12.4) | 0.861 |
| Right coronary artery | 726(39.9) | 231(69.6) | 495(33.3) | <0.001 | 299(41.8) | 82(80.4) | 217(35.3) | <0.001 |
| Total occlusion of culprit vessel<br>n(%)0=No, 1=Yes | 1259(69.2) | 289(87) | 970(65.2) | <0.001 | 482(67.3) | 95(93.1) | 387(63) | <0.001 |
| Using thrombus aspiration devices<br>during procedure n(%)0=No, 1=Yes | 1112(61.1) | 269(81) | 843(56.7) | <0.001 | 448(62.6) | 74(72.6) | 374(60.9) | 0.026 |
| No- reflow n(%)0=No, 1=Yes | 196(10.8) | 66(19.9) | 130(8.7) | <0.001 | 71(9.9) | 16(15.7) | 55(9) | 0.038 |
| Intra-procedural hypotension n(%)<br>0=No, 1=Yes | 184(10.1) | 116(34.9) | 68(4.6) | <0.001 | 60(8.4) | 43(42.2) | 17(2.8) | <0.001 |

In univariable analysis, 10 variables (age, sex, history of coronary artery disease, the culprit vessel was LAD, the culprit vessel was right coronary artery (RCA), the culprit vessel was left circumflex (LCX), total occlusion of culprit vessel, using thrombus aspiration devices during procedure, no reflow, and intra-procedural hypotension) were identified as predictors of periprocedural bradycardia. After application of backward variable selection method, AIC, and BIC, 7 variables (no-reflow, the culprit vessel was LAD, using thrombus aspiration devices during procedure, sex, history of coronary artery disease, total occlusion of culprit vessel, and intra-procedural hypotension) remained as significant independent predictors of periprocedural bradycardia during PPCI. Results were shown in Table 2.

**Table 2.** Predictor of periprocedural bradycardia obtained from multivariable logistic regression models in the development data sets

| Periprocedural bradycardia | Odds Ratio | Std.Err | 95% CI | Coef | Std.Err | 95% CI | Z | P>\| Z \| |
| --- | --- | --- | --- | --- | --- | --- | --- | --- |
| Sex | .485 | .084 | .346 ~ .681 | -.723 | .173 | -1.061 ~ -.385 | -4.19 | <0.001 |
| The culprit vessel was LAD | .145 | .025 | .103 ~ .204 | .403 | .144 | .121 ~ .686 | 2.80 | 0.005 |
| History of coronary artery disease | 1.497 | .216 | 1.129 ~ 1.986 | .554 | .211 | .141 ~ .967 | 2.63 | 0.009 |
| Total occlusion of culprit vessel | 1.741 | .367 | 1.152 ~ 2.632 | -1.931 | .174 | -2.27 ~ -1.591 | -11.12 | <0.001 |
| Using thrombus aspiration devices during procedure | 2.304 | .431 | 1.596 ~ 3.324 | .835 | .187 | .468 ~ 1.201 | 4.46 | <0.001 |
| No-reflow | 2.126 | .419 | 1.445 ~ 3.128 | .755 | .197 | .368 ~ 1.140 | 3.83 | <0.001 |
| Intra-procedural hypotension | 8.985 | 1.707 | 6.192 ~ 13.038 | 2.196 | .190 | 1.823 ~ 2.568 | 11.56 | <0.001 |
| _cons | .142 | .034 | .089 ~ .226 | -1.955 | .239 | -2.42 ~ -1.486 | -8.17 | <0.001 |

According to the above risk factors we can calculate the predicted probability of periprocedural bradycardia using the following formula:P=1/(1+exp(−(−1.955+-.723*S+.554*TOCV+.755*CNR+.835*TA+-1.93*LAD+.403*CADH+2.196*IH))).CADH= history of coronary artery disease(0=No, 1=Yes),TOCV=total occlusion of culprit vessel(0=No, 1=Yes), CNR=no-reflow(0=No,1=Yes), IH=intra-procedural hypotension(0=No, 1=Yes),LAD=the culprit vessel was LAD(0=No, 1=Yes), S=sex(women coded as 0 and men as 1), TA= using thrombus aspiration devices during procedure (0=No, 1=Yes).

The ROC curve was drawn. AUC was 0.8384 ± 0.0122, 95% confidence interval(CI)= 0.81460∼ 0.86225.

We constructed the nomogram(Figure 1)using the development database based on the 7 independent prognostic markers: no-reflow, the culprit vessel was LAD, using thrombus aspiration devices during procedure, sex, history of coronary artery disease, total occlusion of culprit vessel, and intra-procedural hypotension.

**Figure 1.**
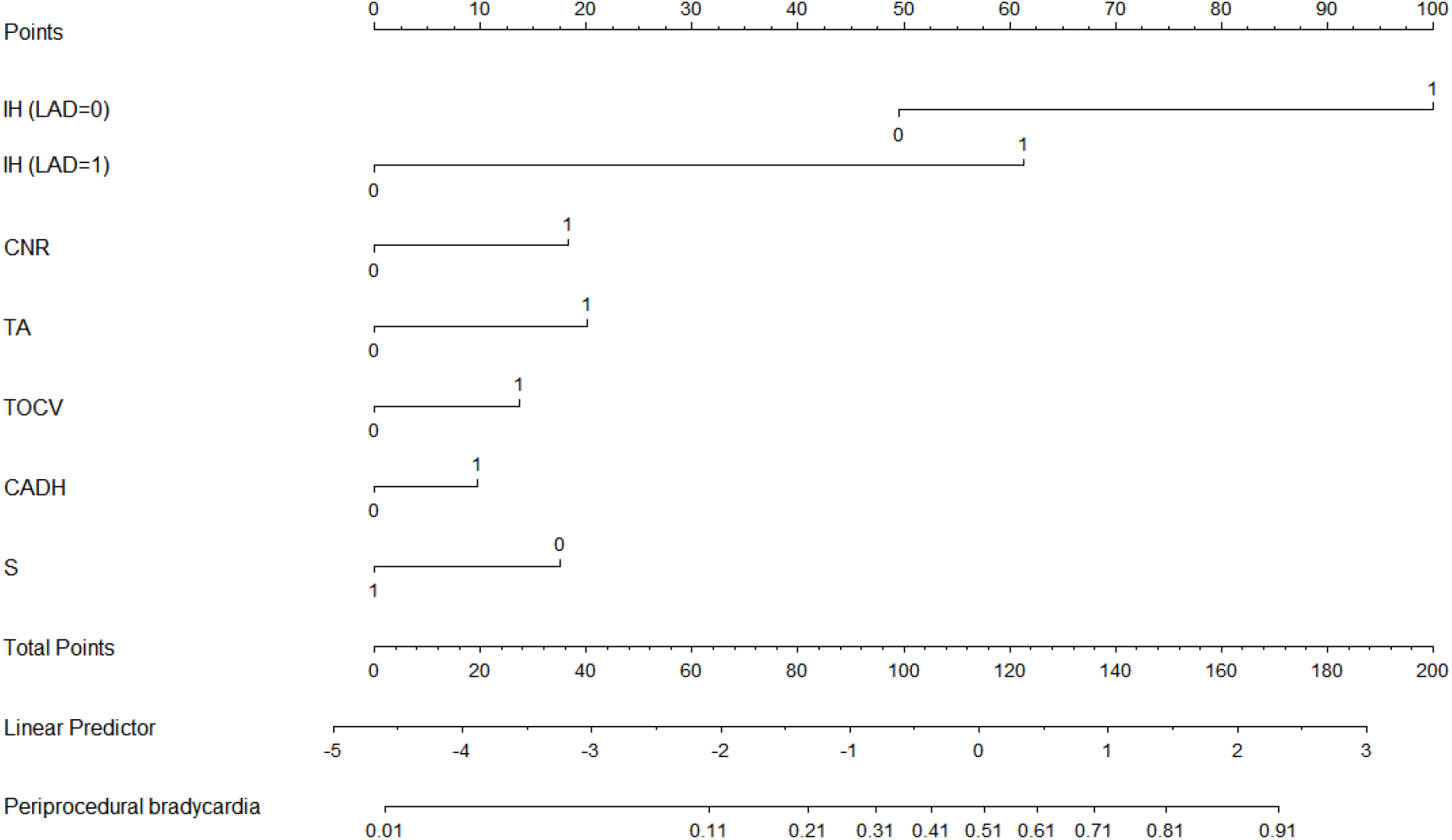
A nomograms for predicting periprocedural bradycardia during PPCI in patients with acute STEMI. LAD =the culprit vessel was LAD(0=No, 1=Yes); CADH = history of coronary artery disease(0=No, 1=Yes); TOCV=total occlusion of culprit vessel(0=No, 1=Yes); CNR = no-reflow(0=No, 1=Yes); IH=intra-procedural hypotension(0=No, 1=Yes); S=sex (women coded as 0 and men as 1); TA=Using thrombus aspiration devices during procedure(0=No, 1=Yes).

Totally 102 patients had periprocedural bradycardia (periprocedural bradycardia group) and 614 patients had no periprocedural bradycardia(control group) in the validation data sets. Baseline characteristics of the patients were shown in Table 1.

We can calculate the predicted probability of periprocedural bradycardia using the following formula:P=1/(1+exp(−(−1.955+-.723*S+.554*TOCV+.755*CNR+.835*TA+-1.93*LAD+.403*CADH+2.196*IH))).CADH=history of coronary artery disease(0=No, 1=Yes), TOCV=total occlusion of culprit vessel(0=No, 1=Yes), CNR=no-reflow(0=No, 1=Yes), IH= intra-procedural hypotension(0=No, 1=Yes), LAD=the culprit vessel was LAD(0=No, 1=Yes), S= sex(women coded as 0 and men as 1), TA= using thrombus aspiration devices during procedure (0=No, 1=Yes).

The ROC curve was drawn. AUC was 0.8437 ±0.0203, 95% CI= 0.80390 ∼ 0.88357.

We drew a calibration plot (Figure2) with distribution of the predicted probabilities for individuals with and without the periprocedural bradycardia in the validation data sets. Hosmer-Lemeshow chi2(10) =15.57,Prob > chi2 = 0.1125>0.05. Brier score=0.0904. DCA in the validation data sets was satisfactory(Figure3).

**Figure 2.**
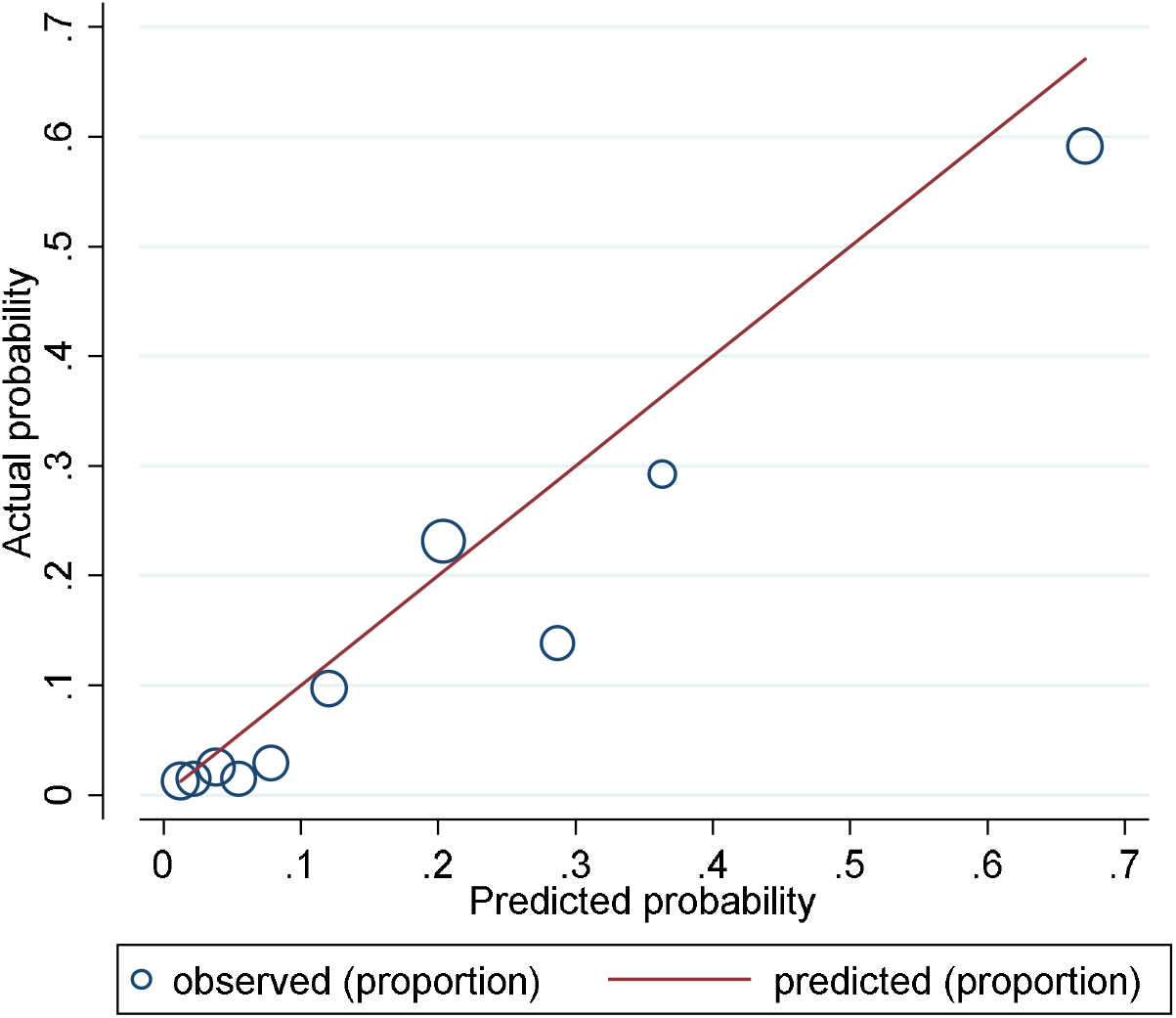
A calibration plot with distribution of the predicted probabilities for individuals with and without the periprocedural bradycardia in the validation data sets.

**Figure 3.**
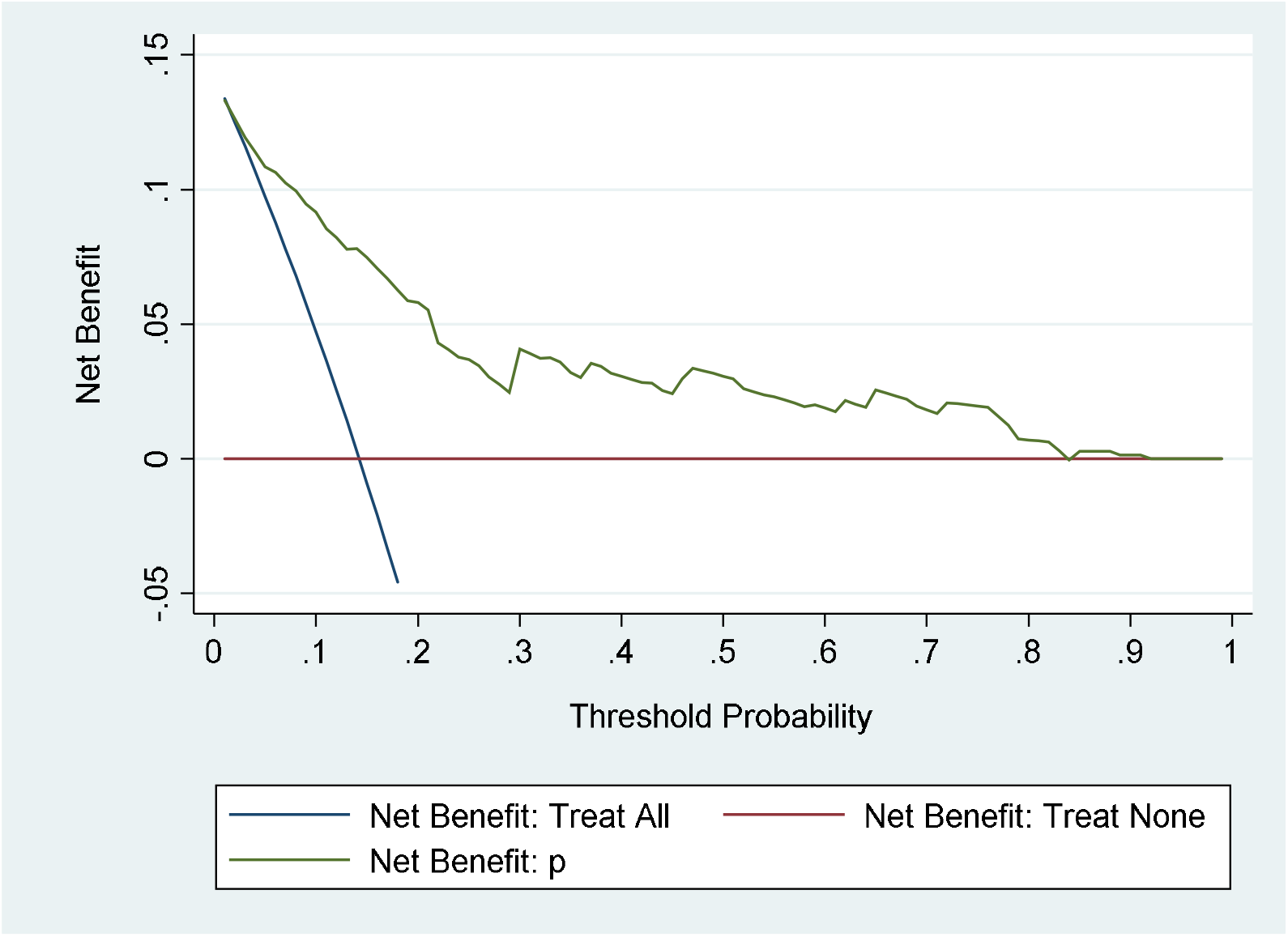
DCA in the validation data sets.

## Discussion

In this study, we investigated the predisposing factors of periprocedural bradycardia in patients with acute STEMI underwent PPCI. A frequency of periprocedural bradycardia was 18.2% (332/1820) in the development data sets. No-reflow, the culprit vessel was not LAD, using thrombus aspiration devices during procedure, sex, history of coronary artery disease, total occlusion of culprit vessel, and intra-procedural hypotension were independent risk factors predicting periprocedural bradycardia during PPCI. ^[11]^ In our study, patients with intra-procedural hypotension were at 8.99 higher risk of periprocedural bradycardia than patients without intra-procedural hypotension.

We assessed the predictive performance of the diagnostic model in the validation data sets by examining measures of discrimination, calibration, and DCA. AUC was 0.8437 ±0.0203, 95% CI=0.80390∼0.88357 in the validation data sets.Prob>chi2=0.1125>0.05.Brier score=0.0904<0.25. Discrimination, calibration, and DCA were satisfactory. We can use the formula or nomogram to predict periprocedural bradycardia. Our diagnostic model of periprocedural bradycardia is not a relative value but an absolute value. The nomogram we constructed for periprocedural bradycardia captures the majority of diagnostic information offered by a full logistic regression model and was more readily used at the bedside.

We followed the discussion of Li et al. 2019. ^[2]^

## Study Limitations

This was a single center experience. Some patients were enrolled >10 years ago thus their treatment may not conform to current standards and techniques. A predictor was more useful when future actions lead to a different proceeding. No-reflow and intra-procedural hypotension were findings which would not alter the interventionalist’s approach too much who was already “in” the situation of re-establishing flow into the vessel.

## Conclusions

We developed and externally validated a diagnostic model of periprocedural bradycardia during PPCI.

## Data Availability

The data used to support the findings of this study are included within the supplementary information file(s).
Code availability (software application or custom code)
The data was demographic, clinical, and angiographic of patients with acute STEMI. AGE=age; CADH=history of corony artery disease; CNR=no-reflow; HD= history of diabetes; HH = history of hypertension; IH=intra-procedural hypotension; LAD=the culprit vessel was left anterior descending; LCX=the culprit vessel was left circumflex coronary artery; PB=periprocedural bradycardia; RCA= the culprit vessel was right coronary artery; S= sex; TA= using thrombus aspiration devices during procedure.

https://pan.baidu.com/s/1rtdC2HIiCXJ_e25d20OO7A

## List of abbreviations

AIC: Akaike information criterion
AMI: acute myocardial infarction
AUC: area under the receiver operating characteristic (ROC) curve
BIC: Bayesian information criterion
CI: confidence interval
DCA: calibration and decision curve analysis
LAD: left anterior descending
LCX: left circumflex coronary artery
PPCI: primary percutaneous coronary intervention
RCA: right coronary artery
ROC: receiver operating characteristic
STEMI: ST elevation myocardial infarction
TIMI: Thrombolysis In Myocardial Infarction flow grade
TRIPOD: Transparent Reporting of a multivariable prediction model for Individual Prognosis Or Diagnosis

## Declarations

### Ethics approval and consent to participate

The study was approved by ethic committee.Approved No. of ethic committee: 2019012X. Name of the ethic committee:Ethics committee of Beijing anzhen hospital capital medical university. It was a retrospective analysis and informed consent was waived by Ethics Committee of Beijing Anzhen Hospital Capital Medical University.

### Statement of human and animal rights

All procedures performed in studies involving human participants were in accordance with the ethical standards of the institutional and/or national research committee and with the 1964 Helsinki declaration and its later amendments or comparable ethical standards. The study was not conducted with animals.

### Consent for Publication

None.

### Availability of data and material

The data used to support the findings of this study are included within the supplementary information file(s).

### Code availability (software application or custom code)

The data was demographic, clinical, and angiographic of patients with acute STEMI. AGE=age; CADH=history of corony artery disease; CNR=no-reflow; HD= history of diabetes; HH = history of hypertension; IH=intra-procedural hypotension; LAD=the culprit vessel was left anterior descending; LCX=the culprit vessel was left circumflex coronary artery; PB=periprocedural bradycardia; RCA= the culprit vessel was right coronary artery; S= sex; TA= using thrombus aspiration devices during procedure.

### Authors’ contributions

Yong Li contributed to generating, analysing, and interpreting the study data and drafted the manuscript. Shuzheng Lyu contributed what to the planning and revised the manuscript critically for important intellectual content. All authors reviewed the manuscript.

Yong Li is being responsible for the overall content as guarantor.

## Acknowledgements

None.

